# Prevalence of depression and its association with health-related quality of life in people with heart failure in low- and middle-income countries: a protocol for systematic review

**DOI:** 10.1101/2023.01.20.23284815

**Authors:** Henok Mulugeta, Peter M. Sinclair, Amanda Wilson

## Abstract

**Background:** Heart failure is a serious clinical burden with variety of physical and emotional symptoms such as fatigue, reduced functional capacity, edema, dyspnea and depression. These symptoms limit patients’ daily physical and social activities, which reduce their health-related quality of life. The objective of this systematic review is to estimate the prevalence of depression and its association with HRQoL in people living with heart failure in LMICs.

**Methods:** The primary outcome is the prevalence of depression in people with heart failure. The secondary outcome is association of depression with health-related quality of life. Comprehensive search of MEDLINE, PsycINFO, EMBASE, CINAHL, Web of Science, Scopus and JBI EBP databases will be conducted to identify relevant studies. The methodological quality of each article will be assessed using a JBI critical appraisal instruments. A random-effects model using the DerSimonian and Laird method will be employed to estimate the regional prevalence. Heterogeneity across the studies will be assessed by Cochrane Q test and I^2^ statistic. A funnel plot and Egger’ s test will be used for assessing publication bias. This protocol is developed in accordance with the JBI methodology for systematic reviews. All statistical analyses will be performed using STATA version 17 software. The Preferred Reporting Items for Systematic Reviews and Meta-analyses (PRISMA) guidelines 2020 will be followed for reporting the results.

**Discussion:** This systematic review will provide up-to-date high-quality evidence on the impact of depression and inform healthcare policymakers on effective ways to improve care for this population in LMICs. The results will be published in a peer-reviewed journal.

**Systematic review registration:** PROSPERO CRD42022361759.

## Background

Cardiovascular diseases (CVDs) are a major global public health problem that are linked to high morbidity and mortality(1). Heart failure is the most common type of CVD in which people experience a variety of symptoms affecting health-related quality of life (HRQOL) (2). Heart failure affects over 64 million people worldwide (3) and was the primary cause of over 300,000 deaths annually in 2019 (4, 5). The incidence of heart failure has increased over the last 10 years due to an ageing population and precipitating factors such as ischemic heart diseases, hypertension and infection (6, 7). Heart failure is a leading cause of morbidity and mortality, with substantial impact on the health care system, due to high readmission rates and associated costs (8).

The burden of heart failure has increased over the past decade in lower and middle income countries (LMICs) with an associated economic and social impact (9). People with heart failure have a significantly higher prevalence of depression than the general population (10). This may be due to comorbidities, polypharmacy, pain, decreased functional capacity, fear of death or disability, and financial problems associated with the disease process (11-13). Depression as a comorbidity of heart failure has been linked to increased readmission, higher health care costs (14-16) and a mortality rate two to three times higher than the general population (17).

Comorbid depression and the inability to carry out daily activities due to the physical symptoms of the disease drastically reduces health related quality of life (HRQoL), and is associated with poor long-term prognoses (18, 19). Adults with heart failure who also have depression and poor HRQoL, are less likely to adhere to self-care recommendations, leading to (20, 21), increased morbidity and mortality (22). In this population, symptoms of depression are often unrecognized and undertreated which increases negative outcomes including all-cause mortality risk (23).

Although studies have been conducted on the prevalence of depression and level of HRQoL in different LMICs, many are of low quality with methodological inconsistencies and inconclusive results. The aim of this review is to estimate and report the current regional prevalence of depression and its association with HRQoL in people with heart failure in LMICs. Providing regional data will help inform healthcare policymakers about strategies to reduce depression and improve HRQoL in people with heart failure in LMICs.

### Research questions

∘ What is the prevalence of depression in people with heart failure in LMICs?
∘ Is there any association between depression and HRQoL in people with heart failure in LMICs?

### Inclusion criteria Participants (population)

This review will consider studies with participants aged 18 years or older who have a diagnosis of heart failure (using Framingham criteria (24) or Echocardiography) with no restriction in terms of gender, ethnicity, or other similar characteristics.

### Condition

This review will consider studies that evaluate the prevalence of depression and/or association of depression with HRQoL in people with heart failure. For the purpose of this review, heart failure is defined as an inability of the heart to effectively pump blood, as defined by either signs and symptoms based on Framingham criteria or reduced ejection fraction (<40%) (25). Depression is the persistent feeling of unhappiness and lack of interest in daily activity, with symptoms lasting for at least two weeks, based on DSM-5 diagnostic criteria (26). Health-related quality of life is defined as an individual perception of physical, mental, emotional, and social health functioning (27). Low to middle income countries will be defined using the World Bank atlas method (28) based on the stratification of economies using gross national income (GNI) per capita. Low-income countries are those with a GNI per capita of $US1045 or less; lower and upper middle-income economies are those with a GNI per capita between $US1046 and 4095 and 4096 and 12695 and respectively.

### Context

This review will consider studies conducted in the context of low-and-middle income countries (LMICs). For the purpose of this review, a list of all LMICs will be compiled using the World Bank database (28).

### Outcomes

This review will consider studies that report the prevalence of depression and/or its association with health-related quality of life using a psychometrically validated instrument.

### Types of studies

This review will consider observational (cross-sectional, cohort, case-control) studies that report the prevalence of depression and/or its association with HRQoL in people with heart failure.

## Methods

This review will be conducted in accordance with the JBI methodology for systematic reviews (29). This protocol is registered in the PROSPERO online database with the protocol registration number CRD42022361759.

### Search strategy

The search strategy aims to locate both published and unpublished studies. Sources will include electronic databases, conference proceedings, websites, dissertations, and direct contact with the author if required. A preliminary search of MEDLINE (Ovid) and CINAHL (EBSCO) was undertaken in August 2022 to identify articles on the topic. The words in titles and abstracts of relevant articles and the index terms used to describe the articles were analysed and used to inform a full search strategy. The search strategy was developed using the **CoCoPop** (**Co**= Condition, **Co**=Context, **Pop**= Population) model. The databases include MEDLINE, PsycINFO, EMBASE, CINAHL, Web of Science, Scopus and JBI EBP database. Index terms (subject headings) and keywords used for the search strategy will be adapted for each database used. The full search strategy for MEDLINE (Ovid) is provided in Appendix I. Reference lists of all identified relevant studies and previous systematic reviews will be screened to identify additional studies. A search for unpublished studies will be conducted using Google scholar, Mednar, ProQuest and dissertation databases. Only studies published in English will be included, with no restriction on date of publication.

### Study selection

Following the search, all identified citations will be collated and uploaded into EndNote V20 (Clarivate Analytics, PA, USA). After removing duplicates, the main author (HM) will screen all titles and abstracts from the original search for assessment against the predefined inclusion criteria. Potentially relevant studies will be retrieved in full, and their citation details imported into the JBI System for the Unified Management, Assessment and Review of Information (JBI SUMARI) (JBI, Adelaide, Australia) (30). The full text of selected citations will be assessed in detail against the inclusion criteria by two independent reviewers (HM and PS). The systematic review will record and report reasons for excluding papers at the full text that do not meet the inclusion criteria. Any disagreements that arise between the reviewers at each stage of the selection process will be resolved through discussion or in consultation with the third reviewer (AW) in order to reach a consensus. The search results and the study inclusion process will be presented in the final systematic review and reported in accordance with the Preferred Reporting Items for Systematic Reviews and Meta-analyses (PRISMA) guidelines (31).

### Assessment of methodological quality

Two independent reviewers (HM and PS) will critically appraise the eligible studies at the study level for methodological quality in the review using a standardised JBI critical appraisal instrument for studies reporting prevalence data (32). The tool is comprised of 9 items that focus on target population, sample size adequacy, study subject and setting (context), reliability of condition measurement, appropriateness of the statistical test used to analyse the data, and adequacy of the response rate with the option to answer “No”, “Yes”, or “unclear”. Authors will be contacted to request missing or additional data for clarification, where required. Following the critical appraisal, the reviewers will either “include” or “exclude” studies based on the overall appraisal quality. A study will be excluded if it has more than three “no” or “unclear” quality categories. This threshold criterion is consistent with that used in similar published systematic review (33). Likewise, the quality of eligible articles to assess the association between depression and HRQoL were also critically appraised using the JBI cross-sectional studies critical appraisal tool for studies reporting association (etiology/risk) (34). Any disagreements will be resolved through discussion. If consensus is not reached, a third reviewer (AW) will be consulted. The results of the critical appraisal will be reported in narrative and table form.

### Data extraction

The JBI data extraction tool for prevalence data and association (etiology/risk) studies will be used to extract pertinent information from each included study. The following information will be extracted from the included studies: authors, year of publication, study date, country of the study, setting, region, design, sample size, outcome (depression), outcome measuring tool, prevalence, and mean score of HRQoL based on exposure (depression), measure of association. When there are missing data, study authors will be contacted and asked to supply relevant information. Two reviewers (HM and AW) will independently conduct the primary data extraction and cross-check for inconsistencies. Any disagreements and discrepancies between the reviewers will be resolved by discussion or with a third reviewer (PS).

### Data synthesis

A narrative presentation of the outcomes (depression and quality of life), including text, table, and figure, will be used to discuss the characteristics of the included studies and synthesise the prevalence of depression and its association with HRQoL. If feasible, data from studies evaluating similar outcomes will be pooled in a statistical meta-analysis using STATA V.14 statistical software. A random-effects model with DerSimonian and Laird model will estimate the pooled effect size of the outcomes, as recommended by Tufanaru et al (35). If there are sufficient data, subgroup analyses will be conducted to investigate the variation between different study characteristics (e.g., region, type of outcome measuring instrument, and study population). Heterogeneity will be assessed statistically using standard chi-squared and I squared tests. The sources of heterogeneity will be analyzed using the subgroup analysis, and meta-regression. If 10 or more studies are included in a meta-analysis, a funnel plot will be generated in STATA Version 17 to assess publication bias. Statistical tests for funnel plot asymmetry (Egger test, Begg test statistics) will be performed where appropriate. Sensitivity analyses will be conducted to test decisions made regarding the effect of a single study on the pooled estimate. The pooled effect size will be presented using a forest plot.

### Assessing certainty in the findings

The Grading of Recommendations, Assessment, Development and Evaluation (GRADE) approach for grading the certainty of evidence will be followed (36), and a summary of findings will be created using GRADEpro GDT (McMaster University, ON, Canada). The outcomes reported in the summary of findings will be prevalence of depression and mean quality of life score. The certainty of evidence will be rated as high, moderate, low, or very low. Two independent reviewers will undertake this at the outcome level. Any disagreements between the reviewers will be resolved through discussion or with a third reviewer. Authors of papers will be contacted to request missing or additional data for clarification where required.

## Discussion

Heart failure is the most common cardiovascular disease and affects about 26 million people worldwide(37, 38). It is a leading cause of morbidity and mortality, substantially impacting the health care system due to high readmission rates, and has a significant effect on mental health and quality of life (39, 40). The prevalence of self-reported depressive symptoms ranges between 20-40% among people with heart failure (41). Severe depression is a strong predictor of poor health-related quality of life in people with CVD, particularly those with heart failure (42).

A preliminary search of PROSPERO, MEDLINE, the Cochrane Database of Systematic Reviews and the JBI Evidence Synthesis was conducted on in August 2020. No systematic reviews on this topic were identified. There are two recent systematic reviews which focus on the global prevalence of depression and level of HRQoL in adults with heart failure (43, 44). However, these reviews only used three international databases (Web of Science, Scopus, and PubMed) and were not conducted in accordance with the Preferred Reporting Items for Systematic reviews and Meta-Analyses (PRISMA) statement (31). The findings included very few studies of LMICs, particularly in Africa. Another meta-analysis estimated the pooled prevalence of depressive symptoms in people with heart failure but only included studies from China (23). One systematic review investigated the association of depression with all-cause mortality among people with heart failure (HF), but no pooled data were available regarding the effect of depression on HRQoL(45).

This current review differs from the previous systematic reviews as it will include recently published results from studies conducted in LMICs and examine the effect of depression on HRQoL in people with heart failure.

## Data Availability

All the data are available from the corresponding author up on a reasonable request.

## List of abbreviations

CVD: Cardiovascular diseases;
HF: Heart failure;
HRQoL: Health-related quality of life,
JBI: Joanna Briggs Institute;
LMICs: Low and middle income countries;
PRISMA: Preferred Reporting Items for Systematic reviews and Meta-Analyses

## Declarations

### Ethics approval and consent to participate

Not applicable

### Consent for publication

Not applicable

### Author contributions

HM conceived the study and drafted the protocol. PS and AW provided support and guidance on the search strategy and drafting of the protocol. All three authors (HM, PS and AW) will be involved in data extraction, quality assessment, analysis and contributed to the revision of subsequent manuscript drafts. All authors have read and approved the final draft of the manuscript.

## Acknowledgements

We would like to thank Sarah Su (UTS Faculty of Health librarian) for her assistance in developing and refining the search strategy.

## Authors’ information

This review will contribute towards a Higher Degree Research (PhD) from the faculty of health at the University of Technology Sydney (UTS) for the first author HM. HM is a PhD candidate at UTS under the supervision of Professor Amanda Wilson and Associate Professor Peter Sinclair.

---

**Appendix I:** The full search strategy for MEDLINE (via Ovid), conducted on August 2022.

| No | Query | Records retrieved |
| --- | --- | --- |
| 1 | Depression/ or Depression.tw. | 413590 |
| 2 | Quality of life.tw. or "Quality of Life"/ | 391275 |
| 3 | Health-related quality of life.tw. | 51063 |
| 4 | HRQoL.tw. | 19568 |
| 5 | QoL.tw. | 45371 |
| 6 | Heart failure.tw. or Heart Failure/ | 227166 |
| 7 | heart disease*.tw. or Heart Diseases/ | 236293 |
| 8 | cardiac failure.tw. | 12328 |
| 9 | 1 or 2 or 3 or 4 or 5 | 765187 |
| 10 | (afghanistan or albania or algeria or american samoa or angola or argentina or armenia or azerbaijan or bangladesh or belarus or belize or benin or bhutan or bolivia or "bosnia and Herzegovina" or botswana or brazil or bulgaria or burkina faso or burundi or cabo verde or cambodia or cameroon or central african republic or chad or china or colombia or comoros or congo or costa rica or "cote d'ivoire" or cuba or djibouti or dominica* or ecuador or egypt or arab republic or el salvador or equatorial guinea or eritrea or eswatini or ethiopia or fiji or gabon or gambia or georgia or ghana or grenada or guatemala or guinea or guinea bissau or guyana or haiti or honduras or india or indonesia or iran or iraq or jamaica or jordan or kazakhstan or kenya or kiribati or korea or kosovo or kyrgyz republic or lebanon or lesotho or liberia or libya or madagascar or malawi or malaysia or maldives or mali or marshall islands or mauritania or mauritius or mexico or micronesia or moldova or mongolia or montenegro or morocco or mozambique or myanmar or namibia or nepal or nicaragua or niger or nigeria or north macedonia or pakistan or panama or papua new guinea or paraguay or peru or philippines or romania or russia* or rwanda or samoa or "sao tome and principe" or senegal or serbia or sierra leone or solomon islands or somalia or south africa or south sudan or sri lanka or "st. lucia" or "st. vincent and the grenadines" or sudan or suriname or syrian arab republic or tajikistan or tanzania or thailand or timor leste or togo or tonga or tunisia or turkey or turkmenistan or tuvalu or uganda or ukraine or uzbekistan or vanuatu or vietnam or west bank or gaza or yemen or zambia or zimbabwe or low income countr* or middle income countr* or developing countr*).ti,ab,sh,kf. | 1809560 |
| 11 | 6 or 7 or 8 | 446521 |
| 12 | 9 and 10 and 11 | 1230 |
| 13 | limit 12 to "all adult (19 plus years)" | 754 |
| 14 | limit 13 to english | 671 |
| 15 | limit 14 to yr="2012 -Current" | 508 |

**Appendix 2:**
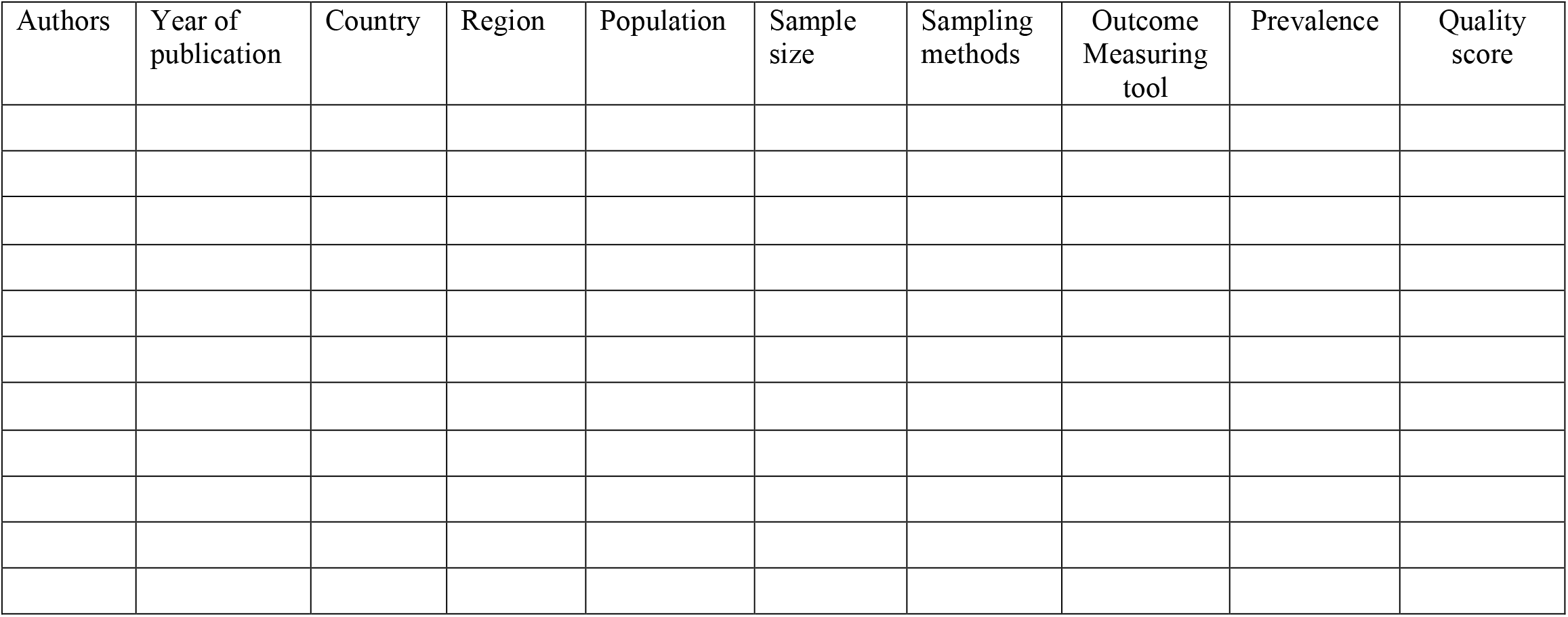
JBI Data extraction form.

